# A meta-analysis of brain DNA methylation across sex, age and Alzheimer’s disease points for accelerated epigenetic aging in neurodegeneration

**DOI:** 10.1101/2020.11.25.20238360

**Authors:** C Pellegrini, C Pirazzini, C Sala, L Sambati, I Yusipov, A Kalyakulina, F Ravaioli, KM Kwiatkowska, DF Durso, M Ivanchencko, D Monti, R Lodi, C Franceschi, P Cortelli, P Garagnani, MG Bacalini

## Abstract

Alzheimer’s disease (AD) is characterized by specific alterations of brain DNA methylation (DNAm) patterns. Age and sex, two major risk factors for AD, are also known to largely affect the epigenetic profiles in the brain, but their contribution to AD-associated DNAm changes has been poorly investigated. In this study we considered publicly available DNAm datasets of 4 brain regions (temporal, frontal, entorhinal cortex and cerebellum) from healthy adult subjects and AD patients, and performed a meta-analysis to identify sex-, age- and AD-associated epigenetic profiles. We showed that DNAm differences between males and females tend to be shared between the 4 brain regions, while aging differently affects cortical regions compared to cerebellum. We found that the proportion of sex-dependent probes whose methylation changes also during aging is higher than expected, but that differences between males and females tend to be maintained, with only few probes showing sex-by-age interaction. We did not find significant overlaps between AD- and sex-associated probes, nor disease-by-sex interaction effects. On the contrary, we found that AD-related epigenetic modifications are significantly enriched in probes whose DNAm changes with age and that there is a high concordance between the direction of changes (hyper or hypo-methylation) in aging and AD, supporting accelerated epigenetic aging in the disease.

In conclusion, we demonstrated that age-associated, but not sex-associated DNAm concurs to the epigenetic deregulation observed in AD, providing new insight on how advanced age enables neurodegeneration.

## INTRODUCTION

Alzheimer’s disease (AD) is a chronic neurodegenerative disease that leads to a progressive decay of cognitive abilities and self-sufficiency. Neuronal loss involves multiple brain regions that are progressively affected by the disease. Hippocampus and entorhinal cortex exhibit the earliest pathological changes, preceding the onset of clinical signs and cognitive impairment by several years, and later the disease spreads to the other brain regions(1-4).

Advanced age and female sex are the two major non-modifiable risk factors for AD(5-7). More than 95% of cases of AD occur after 65 years of age (late onset AD, LOAD), and AD prevalence increases exponentially between 65 and 85 years (8, 9).Two-thirds of clinically diagnosed cases of AD are women, and the fact that women live longer than man does not fully explain this sex bias for AD (10, 11).

The etiology and pathogenesis of AD are complex and likely result from the interplay between genetic and environmental factors during lifespan. In this scenario epigenetic modifications have attracted increased interest in the study of AD, as they integrate genetic background and environment and modulate genomic organization and gene expression. Epigenetic modifications regulate brain biology throughout development and lifetime, influencing neuronal plasticity, cognition and behavior (12), and deregulation of brain epigenetic patterns has been correlated to the pathogenesis of neurological and psychiatric disorders (13, 14). Several studies in post-mortem brain have investigated the role of DNA methylation (DNAm), the best characterized epigenetic modification, in AD, identifying a number of CpG sites that show robust changes in DNAm compared to non-demented controls (15-24).

Interestingly, the two major non-modifiable AD risk factors mentioned above, *i.e*. sex and age, are also among the main biological variables that influence epigenetic patterns in most human tissues, including brain (25).

Genome-wide DNAm differences between males and females have been found in whole blood (26) and have been related to the sex-biased risk of psychiatric diseases (27). A similar link has been reported also in brain (28)where sex-specific DNAm patterns are established early during prenatal development (29, 30)and are at least in part maintained in the adulthood (29, 31), contributing to the profound differences in brain functions between males and females (32-34) and to the different onset of psychiatric disorders(30).

DNAm patterns are largely remodeled during aging (35), where a trend towards global loss of DNA methylation together with hypermethylation at specific loci is observed(36).Although with some differences between brain regions (37, 38), age-associated epigenetic changes interest also the brain, likely contributing to the structural and functional alterations that can resulting progressive cognitive decline and increased susceptibility to neurodegenerative disorders(39, 40).

So far, only few studies have considered how sex and age interact during lifespan in shaping the epigenome. Data on whole blood indicate that sex-dependent DNAm is remodeled during aging (41), and we suggested that these changes occur at different extent in human models of successful and unsuccessful aging. In mouse hippocampus and human frontal cortex, Masser et al identified CpGs in which sex-dependent DNAm is maintained lifelong and CpG sites that are differentially affected by aging in relation to sex (42). Interestingly, some studies employing epigenetic clocks, *i.e*. DNAm-based predictors of age, reported accelerated aging in whole blood from males compared to females (43-45), and the same trend was observed also in brain (43).

Collectively, the available data sustain the importance of sex and aging in shaping the brain epigenome, but so far only one study reported sex-associated DNAm differencesthat were reproducible in different datasets and brain regions (28). No study has systematically analyzed multiple datasets and brain regions to identify DNAm patterns resulting from the interaction of sex and age during lifespan, and most importantly no study has evaluated whether sex- and age-dependent DNAm can contribute to epigenetic deregulation in AD, despite the pivotal role of these two factors in AD etiology and pathogenesis.

To fill this gap, in the present study we performed a meta-analysis of DNAm across sex, age and AD considering publicly available datasets from different brain regions.

## MATERIALS AND METHOD

### Datasets

To select DNA methylation datasets based on Infinium BeadChip technology, the Gene Expression Omnibus (GEO) repository (46) was interrogated by the GEOmetadb Bioconductor package using the following search terms:”GPL13534”, “GPL21145”, to include only datasets based on the Illumina Infinium HumanMethylation450 and MethylationEPIC BeadChips; “sex”, “gender”, “female”, to include only datasets in which the information on the sex of the subjects was available; “age”, to include only datasets in which the information on the age of the subjects was available; “brain”, “cortex”, “gyrus”, “lobe”, “gray”, to select datasets in which brain samples were analysed; “control”, “normal”, “non-tumor”, “health”, or “Alzheiemer”, “AD”, “Braak”, to select datasets including healthy and AD subjects, respectively. We considered only datasets including more than 10 healthy subjects. As to June 30th 2020, only Illumina Infinium HumanMethylation450 datasets were retrieved. As described in the Results section, we applied additional selection criteria according to the different analysis, leaving the datasetsreported in Table 1 and Table 2.

**Table 1.**
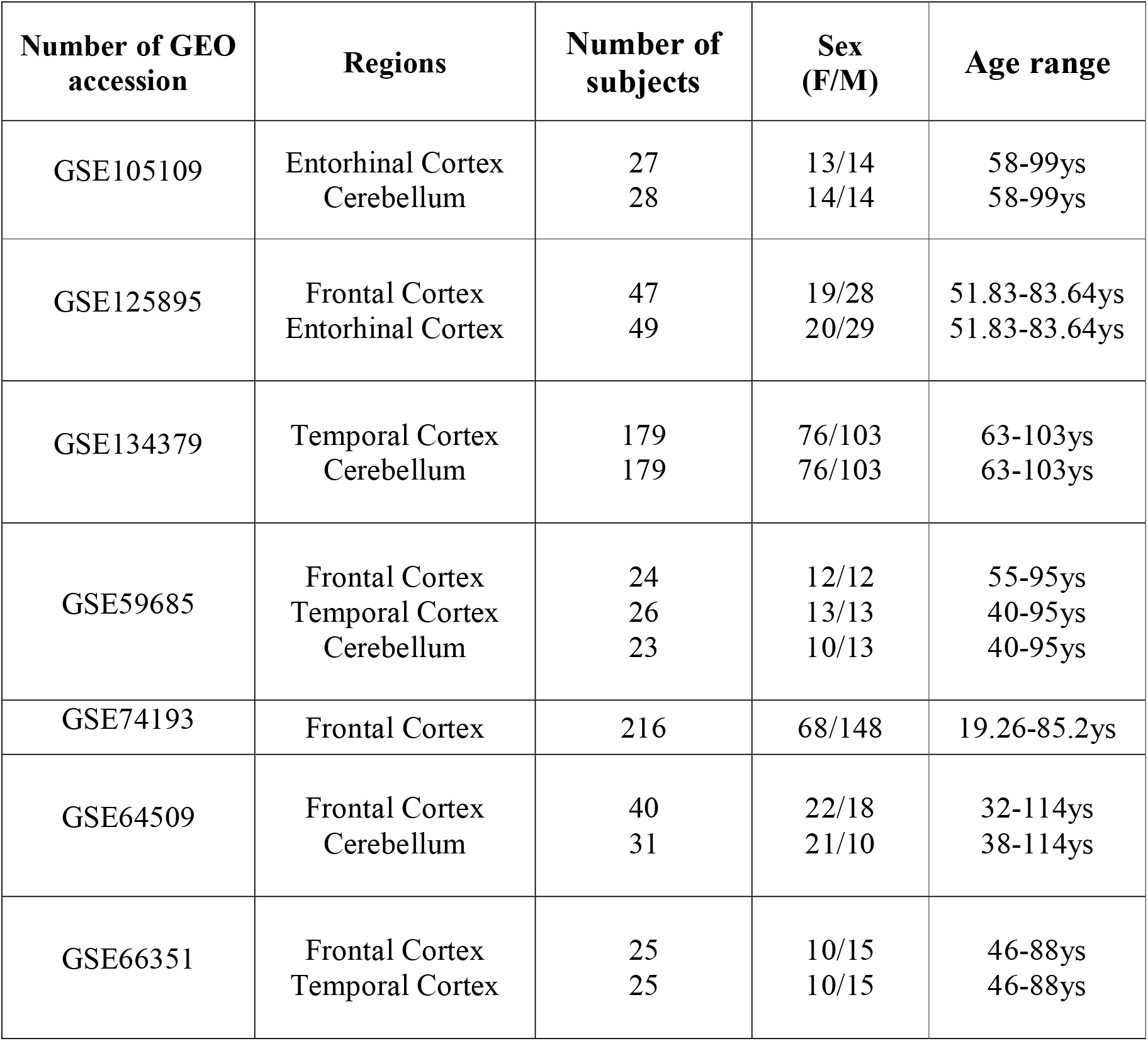
Characteristics of the Infinium450kdatasets including healthy subjects selected in the present study for the age and sex analyses.

| **Number of GEO accession** | **Regions** | **Number of subjects** | **Sex (F/M)** | **Age range** |
| --- | --- | --- | --- | --- |
| GSE105109 | Entorhinal Cortex<br>Cerebellum | 27<br>28 | 13/14<br>14/14 | 58-99ys<br>58-99ys |
| GSE125895 | Frontal Cortex<br>Entorhinal Cortex | 47<br>49 | 19/28<br>20/29 | 51.83-83.64ys<br>51.83-83.64ys |
| GSE134379 | Temporal Cortex<br>Cerebellum | 179<br>179 | 76/103<br>76/103 | 63-103ys<br>63-103ys |
| GSE59685 | Frontal Cortex<br>Temporal Cortex<br>Cerebellum | 24<br>26<br>23 | 12/12<br>13/13<br>10/13 | 55-95ys<br>40-95ys<br>40-95ys |
| GSE74193 | Frontal Cortex | 216 | 68/148 | 19.26-85.2ys |
| GSE64509 | Frontal Cortex<br>Cerebellum | 40<br>31 | 22/18<br>21/10 | 32-114ys<br>38-114ys |
| GSE66351 | Frontal Cortex<br>Temporal Cortex | 25<br>25 | 10/15<br>10/15 | 46-88ys<br>46-88ys |

**Table 2.** Characteristics of the Infinium450kdatasets investigated in the present study including AD patients and non-demented control subjects.

| Number of GEO accession | Regions | Number of subjects | Sex (F/M) | Age range |
| --- | --- | --- | --- | --- |
| GSE105109 | Entorhinal Cortex **Ctrl** | 24 | 13/11 | 66-99ys |
|  | Entorhinal Cortex **AD** | 61 | 27/34 | 67-97ys |
|  | Cerebellum **Ctrl** | 25 | 13/12 | 66-99ys |
|  | Cerebellum **AD** | 64 | 27/37 | 67-97ys |
| GSE125895 | Frontal Cortex **Ctrl** | 11 | 5/6 | 65.04-83.64ys |
|  | Frontal Cortex **AD** | 18 | 9/9 | 71.47-92.29ys |
|  | Entorhinal Cortex **Ctrl** | 12 | 5/7 | 65.04-83.64ys |
|  | Entorhinal Cortex **AD** | 17 | 10/7 | 71.47-92.29ys |
|  | Cerebellum **Ctrl** | 8 | 4/4 | 65.04-83.64ys |
|  | Cerebellum **AD** | 20 | 11/8 | 71.47-92.29ys |
| GSE134379 | Temporal Cortex **Ctrl** | 175 | 76/99 | 68-103ys |
|  | Temporal Cortex **AD** | 217 | 117/100 | 66-102ys |
|  | Cerebellum **Ctrl** | 175 | 74/95 | 68-103ys |
|  | Cerebellum **AD** | 217 | 117/100 | 66-102ys |
| GSE59685 | Frontal Cortex **Ctrl** | 21 | 10/11 | 66-95ys |
|  | Frontal Cortex **AD** | 60 | 39/21 | 66-103ys |
|  | Temporal Cortex **Ctrl** | 22 | 11/11 | 66-95ys |
|  | Temporal Cortex **AD** | 61 | 40/21 | 66-103ys |
|  | Entorhinal Cortex **Ctrl** | 19 | 8/11 | 66-95ys |
|  | Entorhinal Cortex **AD** | 58 | 19/13 | 66-95ys |
|  | Cerebellum**Ctrl** | 19 | 8/11 | 66-95ys |
|  | Cerebellum**AD** | 60 | 39/21 | 66-103ys |
| GSE66351 | Frontal Cortex **Ctrl** | 12 | 8/4 | 71-88ys |
|  | Frontal Cortex **AD** | 35 | 22/13 | 67-97ys |
|  | Temporal Cortex **Ctrl** | 12 | 8/4 | 71-88ys |
|  | Temporal Cortex **AD** | 37 | 23/14 | 67-97y |
| GSE76105 | Temporal Cortex **Ctrl** | 34 | 18/16 | 66-94ys |
|  | Temporal Cortex **AD** | 34 | 17/17 | 66-92ys |
| GSE80970 | Frontal Cortex **Ctrl** | 68 | 34/34 | 70-108ys |
|  | Frontal Cortex **AD** | 74 | 54/30 | 72-103ys |
|  | Temporal Cortex **Ctrl** | 70 | 36/34 | 70-108ys |
|  | Temporal Cortex **AD** | 74 | 54/30 | 72-103ys |
| GSE109627 | Temporal Cortex **Ctrl** | 36 | 19/17 | 73-94ys |
|  | Temporal Cortex **AD** | 46 | 24/22 | 70-95ys |

Ctrl: non-demented control subjects; AD: Alzheimer's disease patients

### Pre-processing

As raw intensities files were not available for some datasets, all the analyses were performed on pre-processed methylation data downloaded from GEO. Probes mapping on sex chromosomes and potentially ambiguous probes (cross-reactive probes and probes including SNPs) (47)were excluded, leaving 415016 probes. In each dataset, neuron/glia proportions were estimated using Horvath’s calculator(48).

### Meta-anaysis

To identify differentially methylated positions (DMPs), the*lmFit* function implemented in *limma* R package (49)was used to fit a linear model to each microarray probe, expressing DNAm as Mvalues. Association with age was calculated using age as a continuous value and correcting for sex and neuron/glia proportion. Association with sex was calculated using sex as a categorical value and correcting for age and neuron/glia proportion. Association with AD was calculated using AD as a categorical value and correcting for age, sex and neuron/glia proportion. The *lmFit* function was used also to calculated the interaction between sex and age, correcting for neuron/glia proportion, and between AD and sex, correcting for age and neuron/glia proportion. Effect sizes and standard errors were extracted from *limma* output.For each brain region, the results obtained in the different datasets were combined by inverse variance-weighted fixed-effects meta-analysis using METAL software (50). Finally, the p-values resulting from each meta-analysis were adjusted for multiple comparisons using the Benjamini-Hochberg (BH) procedure. Only probes with a BH-corrected p-value <0.01 and with concordant effect sizes between all the datasets included in each meta-analysis were retained as significant.

### Enrichment and gene ontology analysis

Enrichment of genomic regions (Islands, N- and S-shores and shelves, open sea regions) was calculated using Fisher exact test, as implemented in the *fisher.test* function implemented in the stats R package. Enrichment of Gene Ontology terms was calculated using the *methylgometh* function implemented in the *methylGSA* R package (51), and redundant GO terms were removed by REViGO software (52).

## RESULTS

### DNA methylation datasets of healthy and AD human brains

We searched GEO database for datasets generated using the Illumina Infinium BeadChips on brain tissues from healthy and AD subjects (Materials and methods).

For the meta-analysis of sex- and age-dependent DNA methylation in healthy subjects, we selected only datasets including at least 10 males and 10 females, having a minimum of 20 years and spanning an age range of at least 30 years. We further considered only brain regions for which at least 2 datasets were available. This resulted in 8 datasets covering 4 regions: Frontal cortex (FC), Temporal cortex (TC), Entorhinal cortex (ERC), Cerebellum (CRB) (**Table 1**).

For the meta-analysis of AD-associated methylation patterns, we selected only the datasets including subjects over 65 years of age with at least 3 males and 3 females in the control and AD groups. This resulted in 8 datasets covering the same brain regions indicating above (**Table 2**).

### DNA methylation differences across sex

To identify sex-dependent differentially methylated positions (sDMPs) we performed an epigenome wide association study (EWAS) in each dataset and brain region separately, correcting for age and estimated neuron/glia proportion (Materials and Methods). We then conducted a meta-analysis within each brain region.

We identified 4860 sDMPs in FC, 1985 sDMPs in TC, 159 sDMPs in ERC and 2322 sDMPs in CRB (**Figure 1A-D, Supplementary Figure 1 and Supplementary File 1**). In FC, sDMPs were mainlyhypermethylated in males compared to females (73% of hypermethylated probes) while the opposite was true for TC,ERC and CRB (38%,33% and 36% of hypermethylated probes in TC,ERC and CRB respectively).When analyzing the genomic context of the sDMPs, we found that CpG islands were enriched in sDMPs in all the 4 brain regions, and that CpG island shores showed a similar trend (**Supplementary File 2**).Also the distribution of sDMPs across chromosomes was not random, with a trend towards enrichment in chromosome 19 in all the 4 brain regions.The enrichment analysis of Gene Ontology (GO) terms did not reveal significant results except for FC, where the “homophilic cell adhesion via plasma membrane adhesion molecules” ontologywas found (**SupplementaryFile 2**).

**Figure 1.**
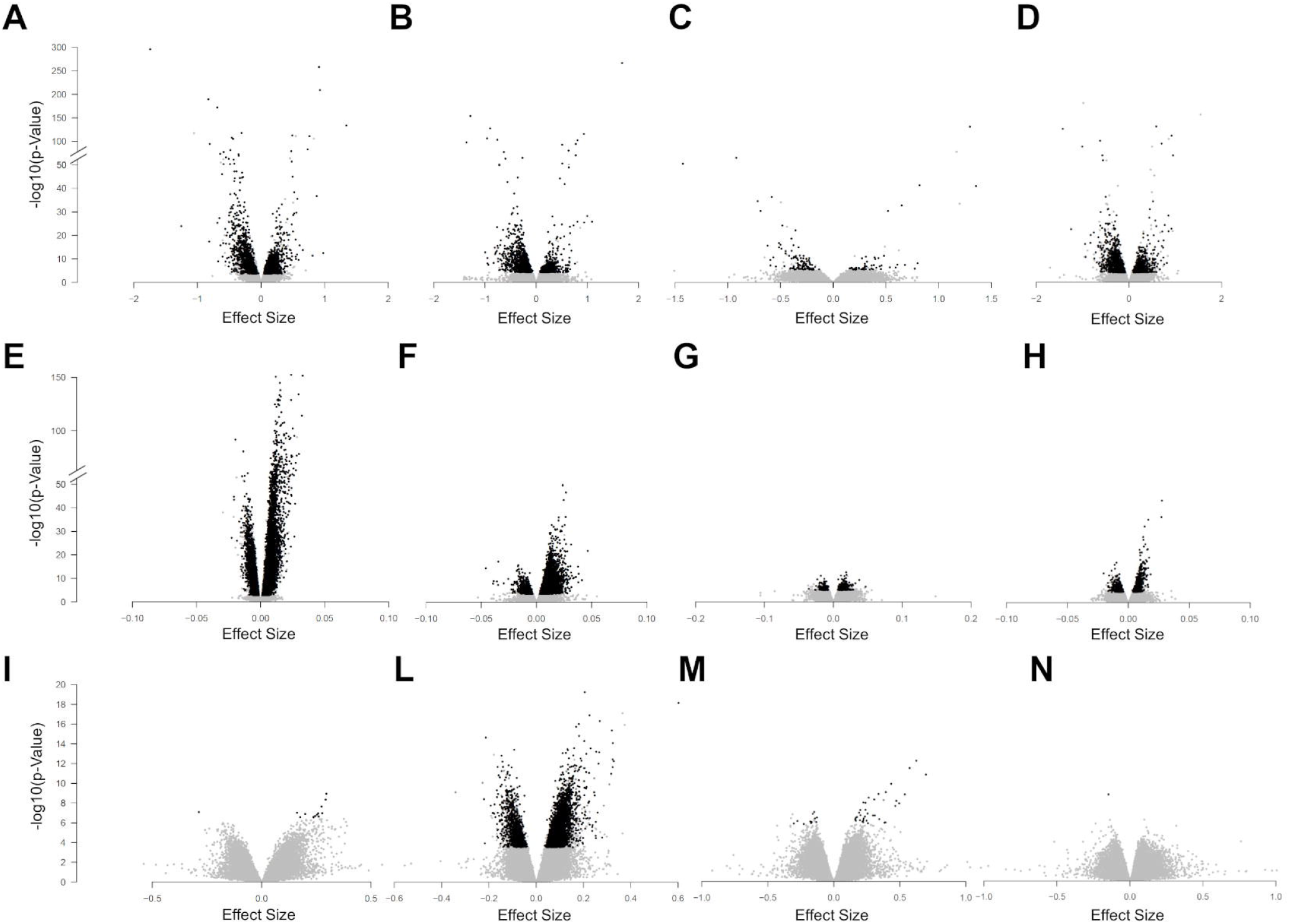
Sex-, age- and AD-associated epigenetic remodeling in the four brain regions. Volcano plots of -log10(P-value) against effect sizes, resulting from the meta-analysis of: sex-associated DMPs in FC **(A)**, TC **(B)**, ERC **(C)** and CRB **(D)**; age-associated DMPs in FC **(E)**, TC **(F)**, ERC **(G)** and CRB **(H)**; LOAD-associated DMPs in FC **(I)**, TC **(L)**, ERC **(M)** and CRB **(N)**. Significant probes (BH-corrected p-value < 0.05) are colored in black.

To investigate whether sex-dependent DNA methylation changes were consistent across brain regions, we evaluated the correlation of effect size values between FC, TC, ERC and CRB (Figure 2A). The 4 brain regions were positively correlated each other (**Figure 2A)**.We next intersected the 4sDMPs lists,identifying77 common probes mapping in 57genes (**Figure 2D, Table 3** and **Supplementary File 1**). All these probes showed concordant sex-dependent DNA methylation profiles in the 4 brain regions and most of them (73%) were hypomethylated in males.

**Table 3.**
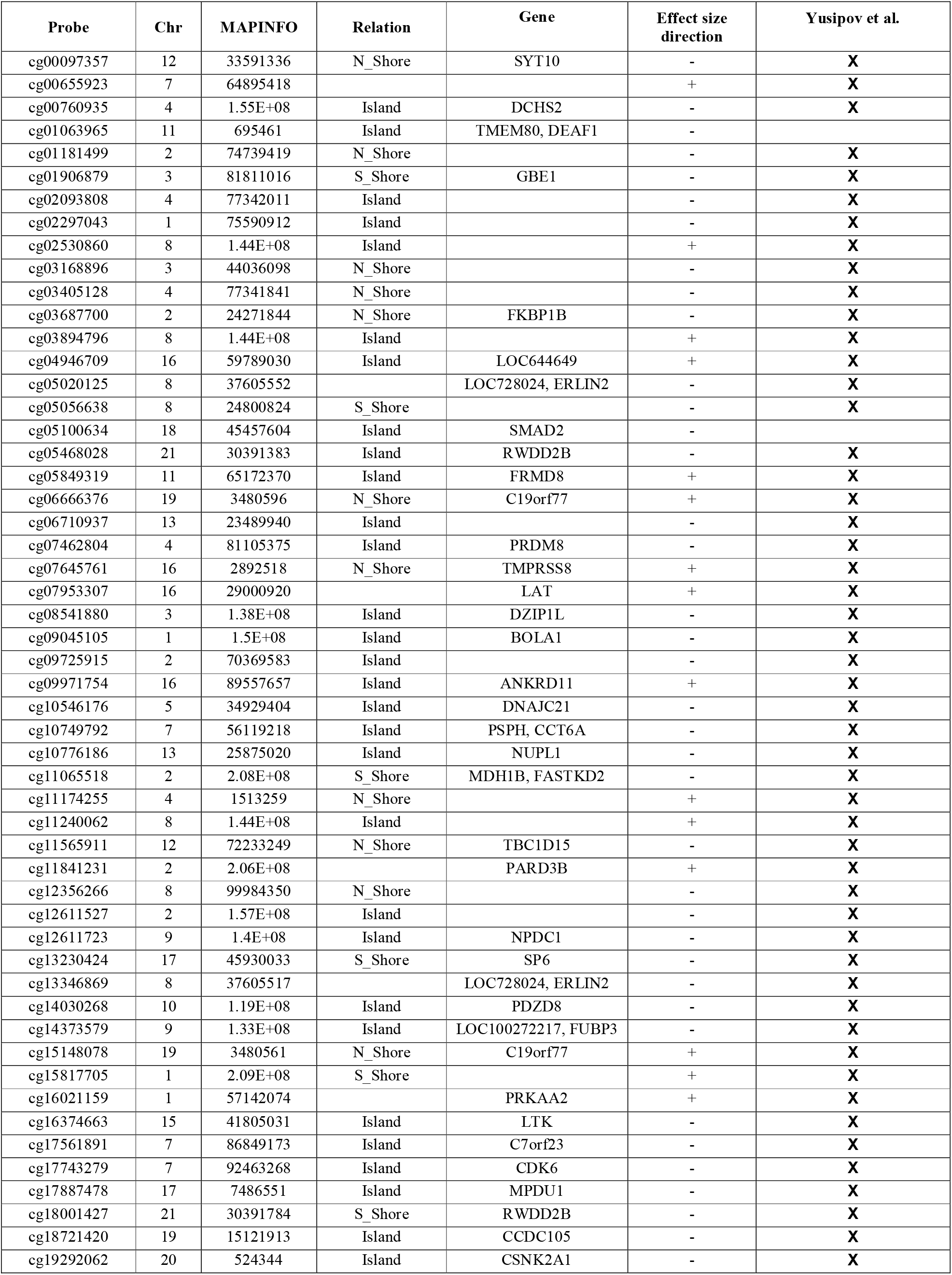
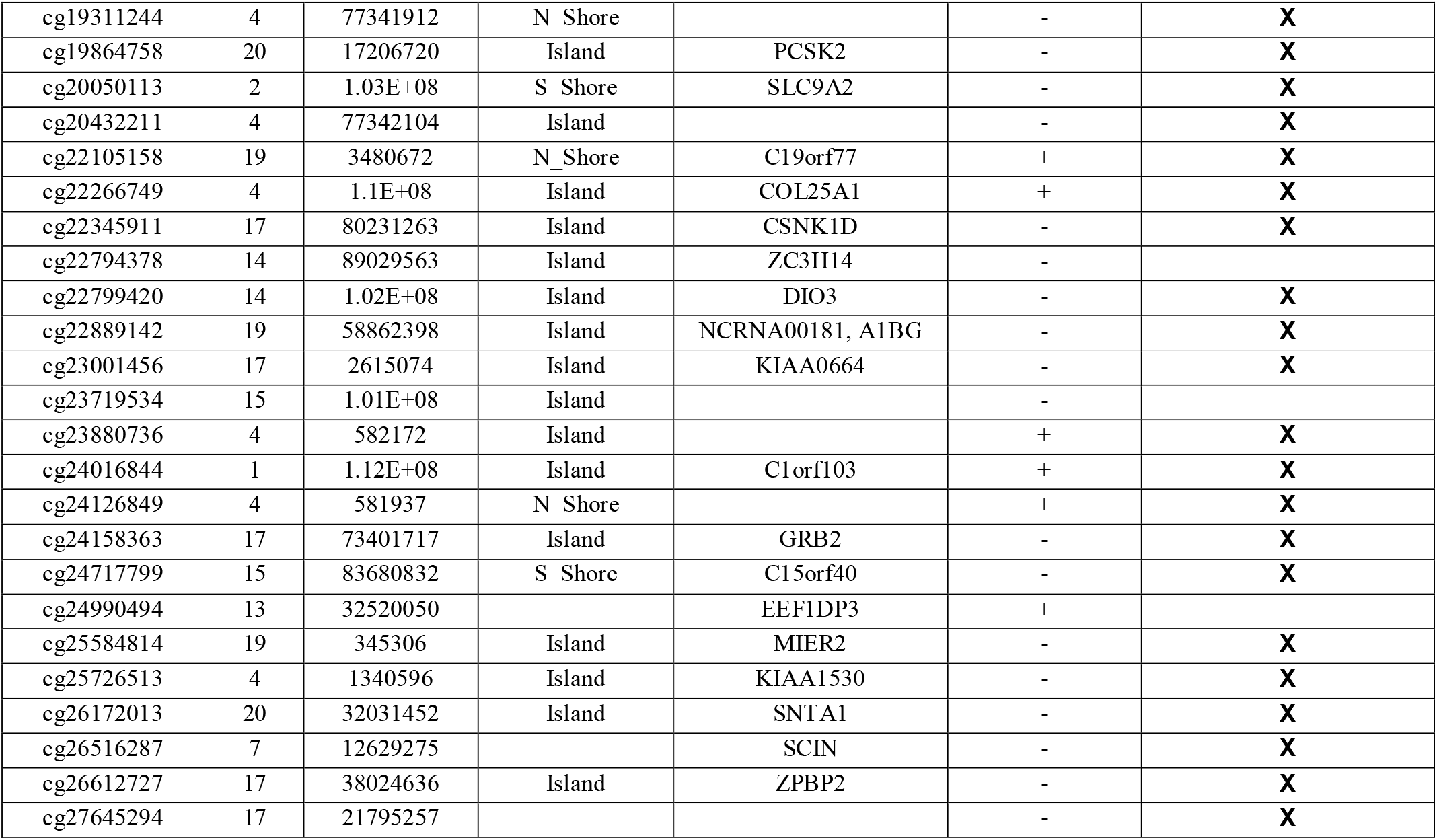
List of sDMPs resulted from cross-region analysis.

**Figure 2.**
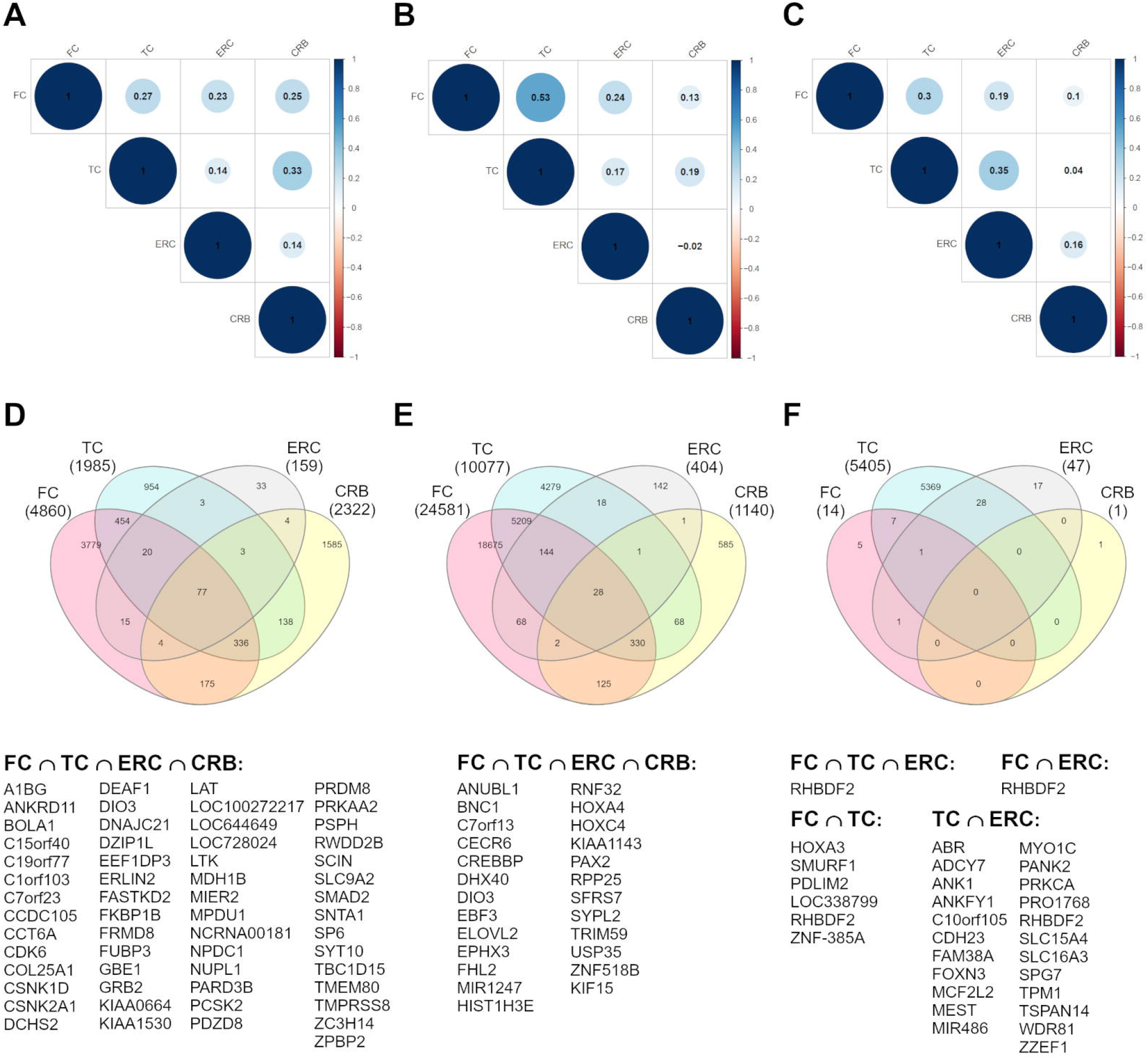
Cross-region analysis of sex-, age- and AD-associated probes. **(A-C)** Correlation matrix plots show the magnitude of correlation between probes effect sizes in the 4 brain regions, considering the result of the meta-analysis onsex- (**A**), age- (**B**) and AD- (**C**) associated probes. Positive and negative correlation values are indicatedin blue and red respectively. **(D-F)** Venn diagrams display the number of significant DMPs shared between the 4 brain regions, considering sDMPs (**D**), aDMPs (**E**) and LOAD-DMPs (**F**). The genes in which map the most shared probes are reported below each diagram.

On the other hand, we searched for probes having sex-related DNAm differences only in one brain region(region-specific sDMPs; Materials and Methods). We found 2, 4, 0 and 37region-specific sDMPs in FC, TC, ERC and CRB respectively (**Supplementary File 1**). Interestingly, 5sDMPs specific for CRB mapped all in the same gene, Nuclear Enriched Abundant Transcript 1 (NEAT1) (**Figure 3**).

**Figure 3.**
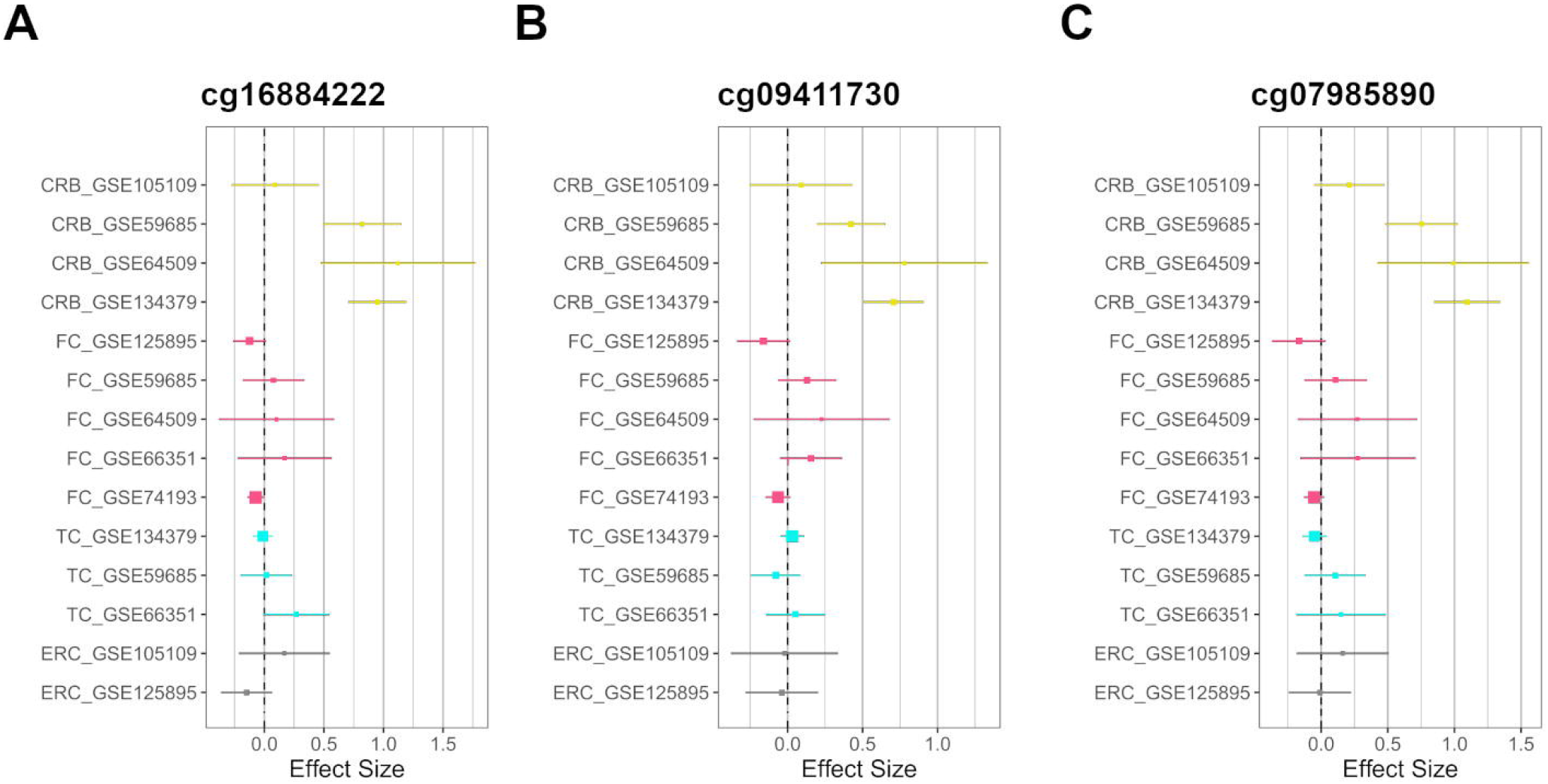
CRB-specific sex-associated DNAm of NEAT1 gene. Forest plot of the three CRB-specific sDMPs mapping in*NEAT1*gene: (**A**) cg16884222, (**B**) cg09411730, (**C**) cg07985890. For each probe, effect sizes from the datasets used for our meta-analysis are reported, dividing them according to the 4 brain regions (CRB, yellow; FC, magenta; TC, cyan; ERC, gray)

### DNA methylation changes across age

To identify age-dependent differentially methylated positions (aDMPs) we performed an EWAS in each dataset and brain region separately, correcting for sex and estimated neuron/glia proportion (Materials and Methods). We then conducted a meta-analysis within each brain region.

We identified 24581, 10077, 404 and 1140 aDMPs in FC, TC, ERC and CRB respectively (**Figures1E-H, Supplementary Figure 2E** and **Supplementary File 3**). In all brain regions, most of theaDMPsunderwent hypermethylation with age (76%, 88%, 58% and 62% of hypermethylatedaDMPs in FC, RC, ERC and CRB respectively). The genomic context of aDMPs was not consistent across the 4 brain regions, except for a significant under-representationin “open sea” regions (**Supplementary File 4**). Similarly, aDMPs were differently scattered across chromosomes in FC, TC, ERC and CRB. GO enrichment analysis revealed several pathways involved in morphogenesis anddevelopmentalprocesses, with “pattern specification process” and “regionalization” common to FC, TC and ERC (**Supplementary File 4**).

The analysis of correlation between the effect sizes revealed that age-associated changes were more similar between FC and TC compared to the other regions (**Figure 2B)**.The intersection of the aDMPs from the 4 brain regions highlighted 28 common probes, all concordantly undergoing hypermethylation with age and mapping in 25 genes (**Figure 2E and Table 4**). The opposite analysis, *i.e*. the identification of region-specific aDMPs (Materials and Methods), identified only 1 probe specific for FC (cg01725130), that maps in the body of Ras And Rab Interactor 3 (RIN3) gene (**Supplementary File 2**).

**Table 4.** List of aDMPs resulting from cross-region

| Probe | Chr | MAPINFO | Relation | Gene | Effect size direction | Yusipov et al. |
| --- | --- | --- | --- | --- | --- | --- |
| cg00292135 | 7 | 156433068 | Island | C7orf13, RNF32 | + | **X** |
| cg04090392 | 15 | 83952774 | Island | BNC1 | + | **X** |
| cg06639320 | 2 | 106015739 | Island | FHL2 | + | **X** |
| cg06942814 | 7 | 27170819 | S_Shore | HOXA4 | + | **X** |
| cg07303143 | 3 | 44803452 | Island | KIAA1143;KIF15 | + | **X** |
| cg07525420 | 10 | 131761181 | Island | EBF3 | + | **X** |
| cg07922606 | 6 | 26225389 | Island | HIST1H3E | + |  |
| cg11614451 | 3 | 160167729 | Island | TRIM59 | + |  |
| cg12373771 | 22 | 17601381 | Island | CECR6 | + | **X** |
| cg13327545 | 10 | 22623548 | Island |  | + | **X** |
| cg14020846 | 14 | 103674272 | Island |  | + | **X** |
| cg14556683 | 19 | 15342982 | Island | EPHX3 | + | **X** |
| cg15243034 | 11 | 77907656 | Island | USP35 | + | **X** |
| cg15341124 | 14 | 102027734 | Island | DIO3, MIR1247 | + | **X** |
| cg15611336 | 15 | 75248496 | Island | RPP25 | + | **X** |
| cg16295725 | 4 | 10459219 | Island | ZNF518B | + | **X** |
| cg16867657 | 6 | 11044877 | Island | ELOVL2 | + | **X** |
| cg16969368 | 17 | 57642752 | Island | DHX40 | + | **X** |
| cg18008766 | 2 | 38978896 | S_Shore | SFRS7 | + | **X** |
| cg18240400 | 10 | 46168597 | Island | ANUBL1 | + | **X** |
| cg18473521 | 12 | 54448265 | S_Shore | HOXC4 | + | **X** |
| cg19399220 | 19 | 10527588 | Island |  | + | **X** |
| cg20591472 | 1 | 110008990 | Island | SYPL2 | + | **X** |
| cg23995914 | 4 | 10459228 | Island | ZNF518B | + | **X** |
| cg24079702 | 2 | 106015771 | Island | FHL2 | + | **X** |
| cg24567591 | 16 | 3931229 | Island | CREBBP | + | **X** |
| cg24903144 | 10 | 102509268 | Island | PAX2 | + | **X** |
| cg26092675 | 6 | 26225258 | N_Shore | HIST1H3E | + | **X** |

### Relation between age and sex in brain DNA methylation

We then aimed at studying how sex-specific brain DNAm is modulated during aging.

First of all, we intersected sDMPs and aDMPslists. In FC, we found 675 probes that change with sex and with age (s&aDMPs), corresponding to about 13% of all sDMPs identified. In TC s&aDMPs were 171, corresponding to 8.5%of sDMPs. In ERC we found only 2s&aDMPs, while in CRBs&aDMPswere 19, corresponding to 4% of sDMPs(**Figure 4 and Supplementary Files 1 and 3**). In all the four regions, the proportion of sDMPs changing with age (i.e., the proportion of s&aDMPs) was higher than expected (Fisher’s Exact Test p-value <0.05; odds ratio of 2.6, 3.8,13.0 and3.0 in FC, TC, ERC and CRB respectively). InFC, TC and CRB, most of the s&aDMPs were probes having higher DNAm levels in males respect to females and undergoing hypermethylation during aging. GO analysis revealed only 1 ontology enriched in FC (“homophilic cell adhesion via plasma membrane adhesion molecules”).

**Figure 4.**
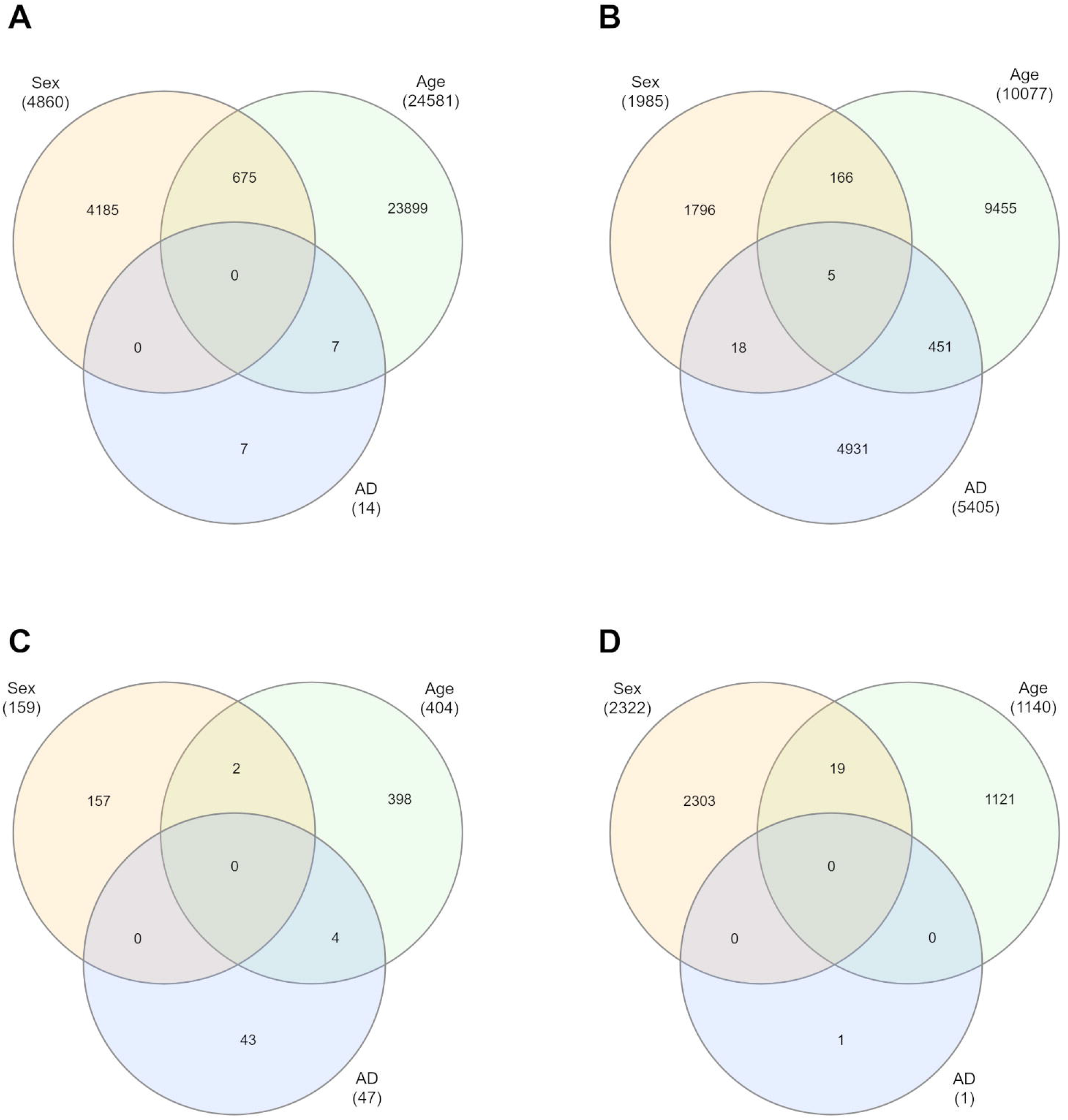
Intersections of sex-, age- and AD-associated probes in each of the four brain regions. Venn diagrams depict the intersection between sDMPs, aDMPs and LOAD-DMPs in FC (**A**), TC (**B**), ERC (**C**) and CRB (**D**).

The previous analysis identifies CpG probes whose DNAm changes according to both sex and age, but is not informative about possible differences in aging trajectories between males and females. To fulfill this point, we performed an age-by-sex interaction analysis in each dataset(Materials and Methods) and meta-analyzed the results for the 4 brain regions. Only 4, 4, 2 and 2 probes showed a significant age-by-sex interaction in FC, TC, ERC and CRB respectively (**Supplementary File 5**).

### Brain DNA methylation changes across AD

Finally, we focused on brain DNAm datasets including late onset AD patients (LOAD) and age-matched non-demented controls.

To identify differentially methylated positions associated with LOAD (LOAD-DMPs) we performed an EWAS in each dataset and brain region separately, correcting for age, sex and estimated neuron/glia proportion (materials and methods). We then conducted a meta-analysis within each brain region.

We identified14 LOAD-DMPs in FC, 5405 in TC, 47 in ERC and only 1 in CRB (**Figures 1I-N**), **Supplementary Figure 3** and **Supplementary File 6**). In all brain regions most of LOAD-DMPs were hypermethylated in AD compared to controls (93%,80%, 76% and 100% in FC, TC, ERC and CRB respectively).While in TC LOAD-DMPs were significantly under-represented in CpG islands and enriched in the other genomic contexts, a significant enrichment in CpG islands was found for LOAD-DMPs identified in FC (**Supplementary File 7**). GO analysis returned significant results only in TC, where pathways related to synapse organization and function were found (**Supplementary File 7**).

Correlation analysis of effect size between 4 brain regions highlighted a distinctive pattern in CRB respect to FC, TC and ERC, while the correlation was higher between TC and ERC (**Figure 2C**). Accordingly the intersection between LOAD-DMPs in the 4 brain regions did not return common probes, while 29 probes (mapping in 23 genes)and 8 probes (mapping in 6 genes) were identified by intersecting TC and ERCor FC and TC, respectively (**Figure 2F**and **Table 5**). The probecg12163800, mapping in Rhomboid 5 Homolog 2 (RHBDF2) gene, was significantly hypermethylated in FC, TC and ERC from AD patients.

**Table 5.**
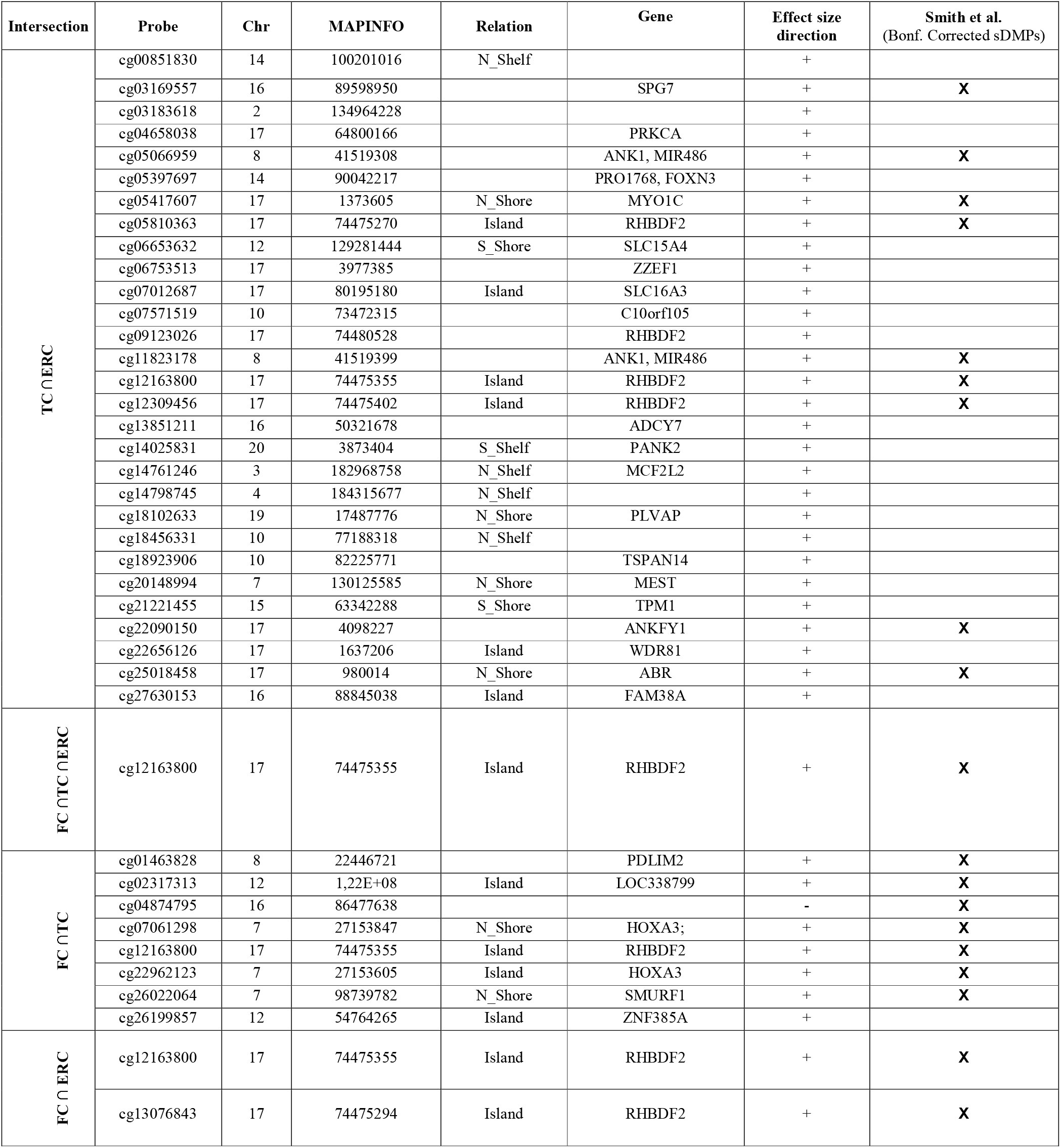
List of LOAD-DMPs resulting from cross-region analysis.

### Relation between sex- and age-associatedDNAm changes and AD epigenetic remodeling

Finally, we explored whether AD-associated DNAm changes were related to sex- and age-specific brain DNAm patterns occurring in physiological conditions, identified in the above described analyses.

In each brain region, we intersected the LOAD-DMPs and sDMPs in orderto identify LOAD&sDMPs, *i.e*. probes that have basal differential DNAm between the two sexes and are also affected by AD.The intersection did not result in any probe for all the regions except that for TC, where we found 23LOAD&sDMPs, mapping in 16 genes and corresponding to only0.4% of LOAD-DMPs in TC (Fisher’s Exact Test p-value >0.05)(**Figure 4**and**Supplementary Files 1 and 6)**.Moreover, AD-by-sex interaction analysis yielded no significant probes in any regions.

Similarly, we explored whether LOAD-DMPs occur in probes whose DNAm changes during physiological aging process (LOAD&aDMPs). The intersection between LOAD-DMPs and aDMPs highlighted 7, 456, 4 and 0 probes in FC, TC, ERC and CRB respectively(**Figure 4**).The proportion of LOAD&aDMPs was higher than expected by chance in FC, TC and ERC (Fisher’s Exact Test p-value <0.05; odds ratio of 15.9, 3.8 and 95in FC, TC and ERC respectively). We found that the 87% of LOAD&aDMPs in TC are concordant for the effect size sign between aDMPs and LOAD-DMPs, while this percentage reached 100% in FC e ERC. Notably, the 4LOAD&aDMPsfound in ERC (cg11823178, cg03169557, cg25018458 and cg22090150, mapping in SPG7 Matrix AAA Peptidase Subunit, Paraplegin [SPG7], Ankyrin1 [ANK1/MIR486], Ankyrin Repeat And FYVE Domain Containing 1 [ANKFY1] and ABR Activator Of RhoGEF And GTPase [ABR] respectively)were also found in TC.Also the intersection between LOAD&aDMPs in TC and FC resulted in 4 common probes: cg22962123, mapping in PDZ And LIM Domain 2 (PDLIM2) gene; cg07061298, not mapping in any gene; cg01463828 and cg04874795, both mapping in Homeobox A3 (HOXA3) gene. Figure 5 reports DNAm values of cg11823178 (ANK1) and cg22962123 (PDLIM2) in TC from GSE134379 dataset as an example of CpG sites displaying a positive association of DNAm with age and hypermethylated in AD.

**Figure 5.**
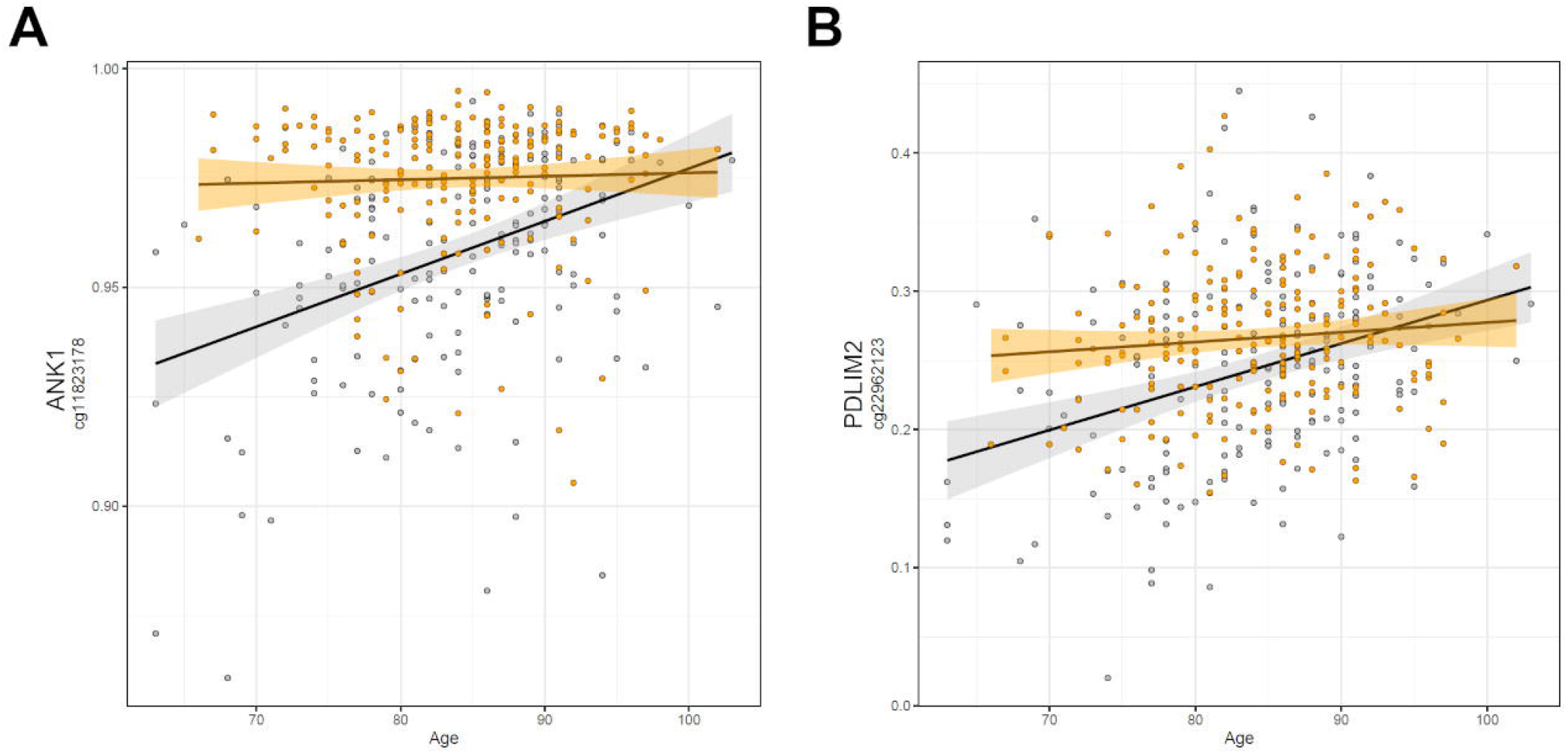
Scatter plots of *ANK1* and *PDLIM2* DNAm according to age and disease. Scatter plots of methylation values of cg11823178 within *ANK1*(**A**) and of cg22962123 within *PDLIM2* (**B**) in TC from GSE134379 dataset). Healthy subjects are colored in gray while AD patients are in orange. Regression lines and interval confidence within each group are reported.

Finally, it is worth to note that TC is the exclusive brain region in which we found probes at the intersection between aDMPs, sDMPs and LOAD-DMPs (LOAD&a&sDMPs)(**Figure 4B**).The 5 probes (cg20225999, cg03951603, cg08820801, cg22263793, cg10828284) which mapped inF-Box Protein 17 (FBXO17), Mov10 Like RISC Complex RNA Helicase 1(MOV10L1)genes were all hypermethylated in males and with aging; three of them (cg20225999, cg08820801, cg10828284) were further hypermethylated in AD.

## DISCUSSION

Sex and age are among the major risk factors for AD. In this paper we performed a meta-analysis of DNAm changes that are associated to sex and aging in 4 brain regions (FC, TC, ERC, CRB) and we evaluated whether they contribute to the epigenetic alterations that have been widely described in AD. Our main findings are summarized in the following paragraphs.

### Sex-dependent DNAm differences tend to be shared between brain regions, with few exceptions

To date some studies have reported DNAm sex differences in human brain, mainly focusing on frontal cortex (29-31, 42) with few exceptions (28). Our meta-analysis confirms the presence of autosomic probes with differential methylation between males and females in all the brain regions. These probes preferentially map in CpG islands and shores suggesting their involvement in the regulation of sex-specific gene expression in brain(31).

Sex specific DNAm tend to be reproducible across the brain regions and 77 CpGs resulted from the cross-region intersection. Among them there are sDMPs mapping in genes that have been already associated to sex differences in brain physiology and pathology, likePar-3 Family Cell Polarity Regulator Beta (PARD3B)(53), DEAF1 Transcription Factor (DEAF1)(54) and Iodothyronine Deiodinase 3 (DIO3)(55) genes. Most of these 77 probes were previously reported as differentially methylated between males and females also in previous meta-analysis on blood(56, 57).

In addition, we found few examples of sDMPs specific for a brain region. The most notable example is in cerebellum and maps in NEAT1. NEAT1 is a ubiquitously expressed long non-coding RNA (lncRNA) involved in a plethora of neurospecific processes such as brain development and aging (58-60). Recent transcriptomic studies on human CNS revealed altered NEAT1 levels in AD (61),PD (62) and in schizophrenia (63).

### DNAm tends to be differently remodeled during aging according to the brain region

Several studies have analyzed age-associated changes in DNAm in brain, both comparing fetal *versus* adult brains and analyzing methylation profiles across adulthood(13, 16, 38, 64-67). Our meta-analysis showes that during aging there is an increase of methylation at specific loci, accordingly to previously published data on blood (56, 68)and brain (38). As previously reported by Hernandez et al (38), also our results support the involvement of brain aDMPs in GO related to developmental processes and morphogenesis. Furthermore, our meta-analysis confirms and extends the observation that the epigenome is differently remodeled during aging across brain regions(38).In particular we observed thatage-associated DNAm patterns are similar in TC and FC, while they are distinct in ERC and CRB. CRB was previously described to undergo to a peculiar epigenetic aging, which was decelerated according to Horvath’s epigenetic clock(37).

The large fraction (93%) of the 28 aDMPs emerged from our cross region analysis was found also in aging studies on blood (56). Among them there are probes mapping in Four And A Half LIM Domains 2 (FHL2) and ELOVL Fatty Acid Elongase 2 (ELOVL2) genes, previously reported as age-associated in large number of studies on several tissues (69, 70)including sorted neuron and glia cells(16). According to what discussed above and to previous results (69, 71) the effect size of ELOVL2 probe cg16867657 was lower in CRB respect the other regions, but still significant in our meta-analysis. Elovl2 is an enzyme involved in the elongation of fatty acids and its functional role in aging has been recently suggested (72).

### Sites with sex-dependent DNAm are similarly modulated during aging in males and females

Previous studies in mice and humans suggested that, while sex-differences in DNAm at certain CpG sites are maintained during life, other CpG sites show sexually divergent aging patterns, *i.e* they have a different response to aging in males and females(42). Our meta-analysis supports the fact that sDMPs have a high propensity to be modulated during aging, as the intersection of sDMPs and aDMPs is higher than expected in all the 4 brain regions. However, we found only few probes with significant sex-by-age interaction, indicating similar rather than diverging changes in DNAm in males and females aging. The discrepancy between our results and previous findingscan be due to different reasons: for example, while Masser et al. considered only one dataset including frontal cortex data, here we meta-analysed several datasets using selective criteria of concordance between all datasets from the same brain region; furthermore, we applied a filtering step that removed potentially ambiguous probes, thus reducing the potential overlap with Masser’s results. Our results are more similar to what reported by two independent studies in blood (41, 56),which showed that only a small fraction of CpGs have significant sex-by-age interaction. Further studies on larger cohorts are needed to better describe sex-dependent DNAm patterns during brain aging.

### Epigenetic changes in AD are enriched in sites that show age-but not sex-dependent DNAm

A recent meta-analysis on EWAS studies identified 220 CpGs associated with AD neuropathology, shared by brain cortical cortex regions but not by CRB(73). The paper by Smith et al. included several datasets that we used also in our meta-analysis, with the exception of GSE125895 and GSE109627, while we did not have access to the ROS/MAP and RBD DNAm data. Furthermore, while Smith et al. considered the association with Braak stage, here we used the disease as a binary trait (affected/unaffected). Despite these differences, our results largely overlap with those previously reported. In particular, we did not find AD-related probes common to all the 4 brain regions that we investigated, with CRB DNAm clearly less affected by the pathology. On the contrary, a subset of sites was shared between FC, TC and ERC, and about 50% of these probes overlap with published data. These probes map within genes whose epigenetic deregulation has been largely documented in AD, including *ANK1, RHBDF2* and *HOXA3*. On the contrary, we did not find any overlapwhen comparing our results on AD brain with CpG sites identified in AD patients’ blood (74), confirming that the pathology differently affects the two tissues as recently reported(75).

We did not find a significant overlap between LOAD-DMPs and sDMPs, nor we found significant interaction effects between sex and AD. Overall these results suggest that AD does not predominantly insist on sites with sex-specific DNAm, and that the epigenetic differences between the two sexes do not contribute to the different prevalence of the disease in males and females.

Conversely, our data show that in FC, TC and ERC, AD-related epigenetic modifications are significantly enriched in probes whose DNAm changes with age. Strikingly, we found a high concordance between the direction of DNAm changes (hyper or hypo-methylation)in LOAD&aDMPs, indicating that a subset of age-associated DNAm changes are exaggerated in AD. In TC, LOAD&aDMPs included probes mapping in*ANK1*, and it is worth to note that the down-regulation of Ank2 (ANK1 human orthologue gene) in *Drosophila* has been associated to memory loss, neuronal dysfunction and shortened lifespan in a recent report(76).

Overall, these results support a geroscience view(77-80)according to which AD can be considered a deviation of the physiological aging trajectories towards accelerated aging. Epigenetic age acceleration was previously reported in AD neurons, were a pronounced loss of CpH methylation was found at enhancers, similar to what observed in aging, and in bulk prefrontal cortex, where epigenetic age calculated by Horvath’s clock was positively associated with neuritic plaques and amyloid load (81). It will be interesting to know whether similar results will be obtained using the recently published epigenetic clock optimized for brain tissues(82).

### Strengths, limitations and conclusion

To the best of our knowledge, this is the first report in which sex-, age- and AD-related DNAm changes are systematically assessed using the same analytical approach. Furthermore, we used stringent selection criteria that enabled to select only probes with concordant DNAm changes in the different datasets. On the other side, our study has some limitations. The datasets that we meta-analysed largely vary in size and age range of the assessed subjects, an aspect important for the identification of aDMPs. Moreover, in all the datasets it was not possible to distinguish 5-methylcytosine from 5-hydroxymethylcytosine, an epigenetic modification that contributes to both brain function and neurodegeneration(17, 73, 83, 84). Finally, the datasets that we meta-analysed were based on bulk brain tissues. Although all the analyses were corrected for neuron/glia proportions predicted from DNAm data, we cannot exclude that the observed sex-, age- and LOAD-associated DNAm changes are at least in part driven by changes in brain cells composition that occur in physiological and pathological conditions. For example, Gasparoni et al reported that ANK1 deregulation in AD is specific for glial cells (16), a finding further supported by gene expression studies (85), and that the epigenetic profiles of neurons and glia are differently modulated during aging. Notwithstanding, our results suggest that (cell-specific) age-associated remodelling of DNAm is not just a confounding factor for the epigenetic deregulation observed in AD, but on the contrary it is the predisposing *milieu* in which AD pathogenetic mechanisms are established.

In conclusion, we demonstrated that age-associated, but not sex-associated DNAm patterns concurs to the epigenetic deregulation observed in AD, providing new insight on how advanced age enables neurodegeneration.

## Data Availability

Datasets used for the meta-analysis study were downloaded from Gene Expression Omnibus public functional genomics data repository.

https://www.ncbi.nlm.nih.gov/geo/

## Abbreviations

DNAm: DNA methylation
AD: Alzheimer’s Disease
DMPs: differentially methylated positions
LOAD: Late Onset Alzheimer’s Disease
EWAS: Epigenome-Wide Association Study
GO: Gene Ontology
sDMPs: sex-associated differentially methylated positions
aDMPs: age-associated differentially methylated positions
s&aDMPs: sex- and age-associated differentially methylated positions
LOAD&aDMPs: Late Onset Alzheimer’s Disease-specific age-associated differentially methylated positions
LOAD&sDMPs: Late Onset Alzheimer’s Disease-specific sex-associated variably methylated positions
LOAD&a&sDMPs: Late Onset Alzheimer’s Disease-specific sex- and age-associated variably methylated positions

## Supplementary Figures

**Supplementary Figure 1. Manhattan plots of sDMPs in the four brain regions**. The figure displays the Manhattan plots resulting from the meta-analysis of sex-associated probes in FC (**A**), TC (**B**), ERC (**C**) and CRB (**D**). Significant sDMPs are marked with dark color. Scale change across 50 is indicated by an axis break.

**Supplementary Figure 2. Manhattan plots of aDMPs in the four brain regions**. The figure displays the Manhattan plots resulting from the meta-analysis of age-associated probes in FC (**A**), TC (**B**), ERC (**C**) and CRB (**D**). Significant aDMPs are marked with dark color. Scale change across 50 is indicated by an axis break.

**Supplementary Figure 3. Manhattan plots of LOAD-DMPs in the four brain regions**. The figure displays the Manhattan plots resulting from the meta-analysis of AD-associated probes in FC (**A**), TC (**B**), ERC (**C**) and CRB (**D**). Significant LOAD-DMPs are marked with dark color.

**Supplementary file 1. Significant sDMPs in the four brain regions**. The tables report the lists of sDMPs for each brain regions (FC, TC, ERC and CRB). Probes resulting from the analysis of cross-region and region-specific sDMPs are indicated by a cross, together with the probes that are in common with aDMPs or LOAD-DMPs found in the same region.

**Supplementary file 2**.**Enrichment analysis of sDMPs**. The tables report: 1) the results of Fisher’s test on genomic distribution of sDMPs for each brain region, considering genomic context and chromosomal location. Significant results (p-value<0.05) are colored in green or red if depleted or enriched respectively. 2) the results of GO pathway enrichment analysis, after REVIGO filtering. Only the significant results (adjusted p-value <0.01) for each brain region are reported.

**Supplementary file 3. Significant aDMPs in the four brain regions**. The tables report the lists of aDMPs for each brain regions (FC, TC, ERC and CRB). Probes resulting from the analysis of cross-region and region-specific sDMPs are indicated by a cross, together with the probes that are in common with sDMPs or LOAD-DMPs found in the same region.

**Supplementary file 4. Enrichment analysis of aDMPs**. The tables report: 1) the results of Fisher’s test on genomic distribution of aDMPs for each brain region, considering genomic context and chromosomal location. Significant results (p-value<0.05) are colored in green or red if depleted or enriched respectively. 2) the results of GO pathway enrichment analysis, after REVIGO filtering. Only the significant results (adjusted p-value <0.01) for each brain region are reported.

**Supplementary file 5. Probes with significant sex-by-age interaction in the four brain regions**. The tables report the lists of probes with significant sex-by-age interaction in each brain region (FC, TC, ERC and CRB).

**Supplementary file 6. Significant LOAD-DMPs in the four brain regions**. The tables report the lists of LOAD-DMPs for each brain regions (FC, TC, ERC and CRB). Probes resulting from the analysis of cross-region and region-specific LOAD-DMPs are indicated by a cross, together with the probes that are in common with sDMPs or aDMPs found in the same region.

**Supplementary file 7**.**Enrichment analysis of LOAD-DMPs**. The tables report: 1) the results of Fisher’s test on genomic distribution of LOAD-DMPs for each brain region, considering genomic context and chromosomal location. Significant results (p-value<0.05) are colored in green or red if depleted or enriched respectively. 2) the results of GO pathway enrichment analysis, after REVIGO filtering. Only the significant results (adjusted p-value <0.01) for each brain region are reported.

