## Supplementary figures and images for "A meta-analysis of brain DNA methylation across sex, age and Alzheimer’s disease points for accelerated epigenetic aging in neurodegeneration"

### Supplementary figure 1

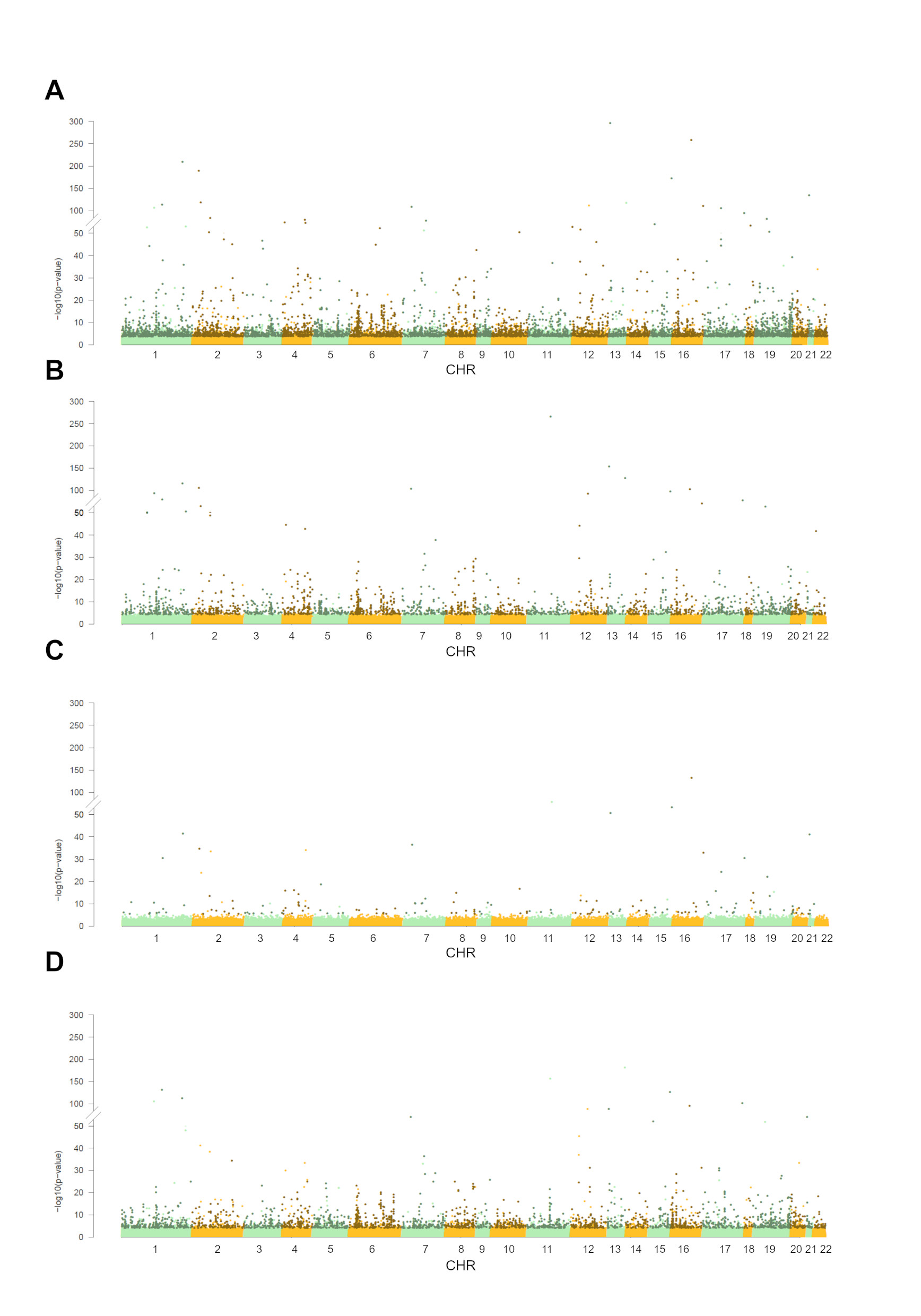

### Supplementary figure 2

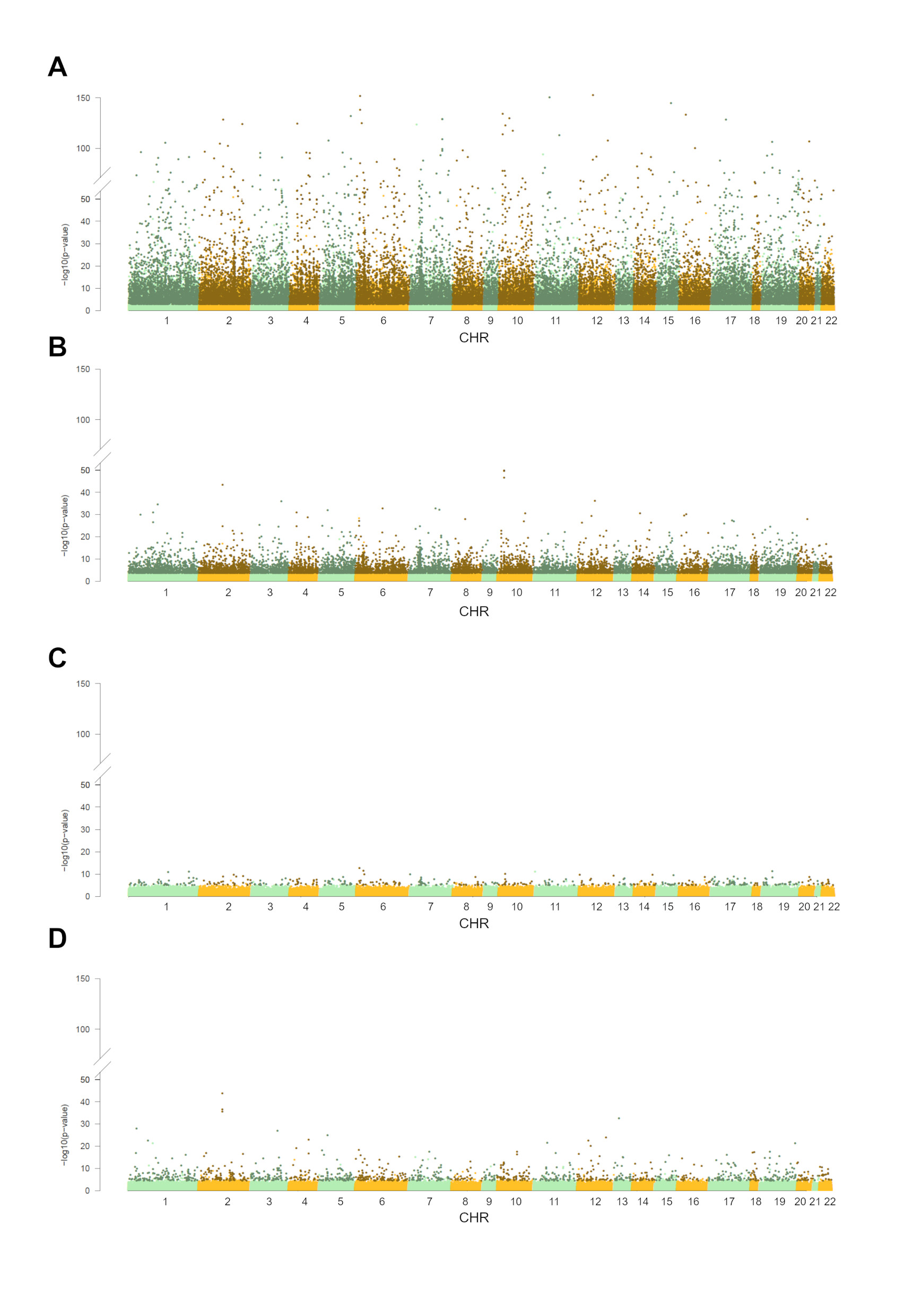

### Supplementary figure 3

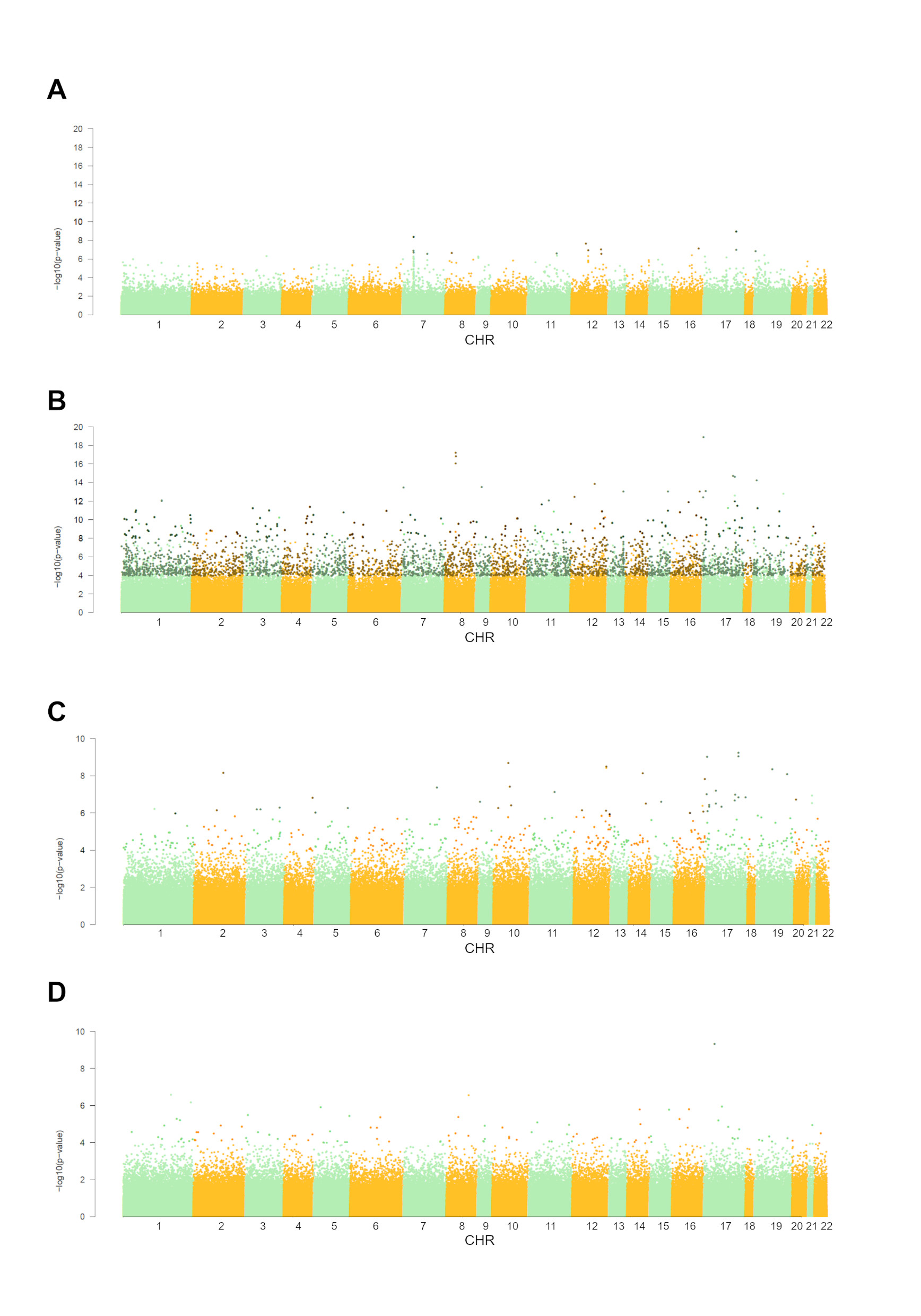
